# Clinical health outcomes of Ebola virus disease survivors eight years post recovery: a cross-sectional study in Sierra Leone

**DOI:** 10.1101/2024.08.29.24312780

**Authors:** Brayden G. Schindell, Boghuma K. Titanji, Anne W. Rimoin, Souradet Y. Shaw, Jia B. Kangbai, Jason Kindrachuk

**Affiliations:** Department of Medical Microbiology and Infectious Diseases, Max Rady College of Medicine, University of Manitoba, Winnipeg, Canada; Division of Infectious Disease, Emory University, Atlanta, United States of America; Department of Epidemiology, Fielding School of Public Health, University of California Los Angeles; Department of Community Health Sciences, Max Rady College of Medicine, University of Manitoba, Winnipeg, Canada; Department of Public Health, Njala University, Sierra Leone

## Abstract

**Background:** The West African Ebola virus disease (EVD) epidemic that occurred between 2013-2016 resulted in >28,000 confirmed cases and >11,000 fatalities. Thousands of survivors necessitate an understanding of the long-term health effects and future medical needs of these patients.

**Methods:** A cross-sectional study of 595 EVD survivors from Sierra Leone and 403 close contacts (*n*=998). An in-person survey conducted between November 2021 and March 2022 included demographics, clinical health symptomology assessment of each organ system and a reproductive health assessment including sexual dysfunction question sets. The frequency of each disorder was examined and the association of each disorder with EVD survival was assessed.

**Results:** Of 12 number of symptom types, five were reported by >50% of EVD survivors (Ocular, Neurological, Constitutional, Genitourinary, Dermatological), and all but one were reported by >40% of EVD survivors. Symptom types associated with EVD survival included ENT symptoms (AOR: 8.75, 95% CI: 5.63 – 13.60, p < 0.001), ocular symptoms (AOR: 7.18, 95% CI: 5.02 – 10.25, p < 0.001), dermatological symptoms (AOR: 4.16, 95% CI: 3.06 – 5.65, p < 0.001) and cardiovascular symptoms (AOR: 2.96, 95% CI: 2.12 – 4.13, p < 0.001).

**Conclusion:** The West Africa EVD epidemic resulted in a high prevalence of persistent health issues among disease survivors. Continued support for survivor services in West Africa is crucial, and future outbreak response planning should include dedicated funding to ensure adequate care for survivors, both during the acute phase of infection and throughout the post recovery period.

## Background

The West African Ebola virus disease (EVD) epidemic of 2013-2016 remains the largest recorded Ebola virus outbreak with 28,652 cases and 11,325 deaths[1]. An estimated 10,000-17,000 individuals recovered from EVD during this period, underscoring the need to address both the immediate impacts of EVD outbreaks and the long-term consequences for survivors. [2]. Sierra Leone, which reported the highest number of cases at 14,124 [1, 3]; has 3,466 registered survivors as of 2020, though estimates suggest the total number could be over 5,000.[2]. Understanding the long-term health effects and future medical needs of survivors is crucial[4]. Most published reports on the health of EVD surivors has focused on the first two years post-recovery, with only two studies extending to four years[5, 6] and one study reaching 7 years. [7].

The “slow and painful” recovery in survivors was first documented in the Sudan EVD outbreak in 1976[8]. More recently the West African EVD epidemic revealed that most EVD survivors experience some measure of persistent health issues[7]. These symptoms and clinical findings manifest as chronic post-infectious syndromes and/or conditions characterized by immune dysfunction[9]. The unprecedented number of survivors has raised questions about long-term health complications of EVD survivorship and the capacity of local health systems to meet these needs[10]. Inconsistent follow-up on the management of sequelae, and limited access to healthcare services have exacerbated the effects of these sequelae[11]. The lack of developed health infrastructure and barriers to accessing care continue to impact the ability of EVD survivors to access the required services that can help improve their overall quality of life[12, 13]. The persistent symptoms experienced by survivors makes it challenging for them to return to their previous lives[14]; for example, most survivors of the Sudan Ebola virus outbreak in the 2000 Gulu, Uganda reported an inability to return to work a year after infection[15].

A wide range of symptoms has been reported among EVD survivors, affecting multiple body systems. The most commonly reported sequelae in the literature include fatigue, joint pain, headache, muscle pain, abdominal pain, ocular complications, anorexia, memory loss, hearing loss and alopecia[3, 4, 6, 7, 14, 16-25]. However, there is ongoing debate regarding the duration of these sequelae, with some studies indicating symptoms wane over time while others suggest they persist[5].

Our study, conducted through a cross-sectional survey, aimed to explore the persistence of sequelae among EVD survivors six to eight years post-survival. We present our findings on the prevalence of sequelae in survivors of the 2013-2016 West African EVD epidemic across Sierra Leone, comparing them to a control group of close contacts from the same regions.

## Methods Study Design

This national cross-sectional study was conducted in Sierra Leone from November 2021 to March 2022. It involved simultaneous recruitment and surveying across all affected districts including ; Bo, Bombali, Falaba, Kailahun, Kambia, Karene, Kenema, Koinadugu, Kono, Moyamba, Port Loko, Pujehun, Tonkolili, Western Area Rural, and Western Area Urban [26]. Given that the estimated national population of EVD survivors in Sierra Leone ranges between 3,000 and 5,000, a sample size of 715 participants was determined to provide 95% confidence intervals for our outcome variables, as previously described for this population[26]. To account for potential non-responses and ensure a minimum sample size for analysis, the recruitment target was increased to 1,000 participants.

### Participants

Eligibility criteria: To qualify as an EVD survivor, individuals were required to provide either an EVD discharge card or a certificate from an Ebola treatment unit in Sierra Leone, dated between March 2014 and March 2016, as proof of surviving the West African EVD outbreak. Eligibility for close contacts required them to verify their relationship to an EVD survivor or a deceased EVD patient. This could be established by presenting an EVD discharge card, a certificate, or a death certificate of the deceased EVD patient relative. Finally, all participants needed to be between the ages of 18 and 50 years at the time of recruitment.

All participants consented to study participation after being fully informed about its scope, potential outcomes, and their right to withdraw at any time without repercussions. Consent was recorded by participants checking the “yes” box in the consent section at the beginning of the survey.

A multistage sampling approach was used, where known survivors provided by the Sierra Leone Association of Ebola Survivors, were stratified by district and gender followed by random sampling within each district, close contacts were identified at the time of recruitment of EVD survivors where survivors were asked if they had any family members interested in participating and had no history of EVD. The multistage sampling started in the South of the country and proceeded counterclockwise ending in the Western region. Due to unreliable internet access in many areas, our recruitment team conducted surveys in person after recruiting participants. Translation to Krio, the most commonly spoken language after English in Sierra Leone was provided, along with assistance to those unable to read.

### Instruments

Socio-demographics were captured as previously described, and included [26] age, sex assigned at birth, district/province of residence, and body mass index (BMI).

Clinical health outcome variables were assessed through symptom screens where participants self-reported symptoms experienced in the past two weeks at the time of the survey. The symptom screen was organized into organ systems, and included chronic medical conditions, infections, ocular symptoms, ear nose and throat (ENT) symptoms, neurological symptoms, constitutional symptoms, cardiovascular symptoms, respiratory symptoms, gastrointestinal symptoms, genitourinary symptoms, musculoskeletal symptoms and dermatology symptoms (a full breakdown can be found in supplemental materials). Participants were asked if they are currently or recently experienced a symptom from a particular organ system and what the frequency of the symptom is, assessed on a 6-point Likert scale from 1 (monthly) to 6 (constantly). Both EVD survivors and close contacts reported symptoms experienced and were also asked whether they had sought medical care and are currently taking medication for symptoms.

Both male and female participants were asked questions on sexual health. Female participants were given the female sexual function index (FSFI) which is a 19-item questionnaire that assesses six dimensions of female sexual health: desire, arousal, lubrication, orgasm, satisfaction and pain[27]. This questionnaire was designed to measure the multidimensional nature of female sexual health. Items were scored on either a 5-point Likert scale from 1 (Very High) to 5 (Very low/none) or a 6-point Likert scale from 0 (No sexual activity) to 5 (Almost always/always) over the last four weeks, with scores ranging from 4 (Severe female sexual dysfunction) to 95 (Normal female sexual function). Female sexual arousal disorder (FSAD) is considered a tentative diagnosis with a score of 57 or less in the FSFI.

Male participants were assessed through the international index of erectile dysfunction (IIEF), which is a 15-item questionnaire that assesses five different domains of male sexual health: erectile function, orgasmic function, sexual desire, intercourse satisfaction and overall satisfaction[28]. Items were scored on either a 5-point [1 (Very High) to 5 (Very low/none)] or a 6-point Likert scale from 0 (No sexual activity) to 5 (Almost always/always) over the last four weeks, with scores ranging from 6 (Severe male sexual dysfunction) to 75 (Normal male sexual function). screening for male sexual dysfunction is considered when scores are 36 or less on the IIEF and screening for erectile dysfunction is considered with a score of 13 or less on the erectile function question subset.

### Statistical Analysis

All statistical analyses in this study were conducted using Stata version 17·0 (College Station, TX: StataCorp LLC; 2021). Descriptive statistics included means and standard deviation, medians and interquartile ranges (IQR), and frequency and percentages, where appropriate. To examine the relationships between independent (e.g. demographic, health-related variables) and dependent variables, either the Chi-square test or Fisher’s exact test was applied. For univariate, bivariate and multivariate analyses were converted into binary variables based on their respective cut-off scores. In bivariate analysis, variables that demonstrated a p-value of 0.1 or lower (using either Chi-square or Fischer’s exact test) were considered for the multivariable logistic regression models. A priori, age, sex assigned at birth, province of residence, and BMI were selected to control for associations between symptomologies and EVD survival status. For the purposes of this study, statistical significance in regression models was defined as a two-tailed p-value of 0.05 or less. Results are presented in the form of adjusted odds ratios (AORs) along with associated 95% confidence intervals (CI).

### Ethical considerations

Ethical approval for this study was obtained from the University of Manitoba Research Ethics Board [HS24515 (H2020:538)] and the Office of the Sierra Leone Ethics and Scientific Review Committee, Government of Sierra Leone.

## Results

### Participant characteristics

A total of 998 participants comprising 403 close contacts and 595 EVD survivors were included, with 533 (53.4%) male participants and 465 (46.6%) female participants[26]. Select participant demographics are described in Table 1; briefly, the average age of participants was not statistically different (p = 0.327) between close contacts (mean = 32.6 years, SD = 8.2 years) and EVD survivors (mean = 32.1 years, SD = 8.9 years). As far as general health of participants which was assessed through body mass index (BMI), there was not a significant difference (p = 0.320) between close contacts (mean = 20.8, SD = 12.1) and EVD survivors (mean = 20.2, SD = 8.0).

**Table 1:**
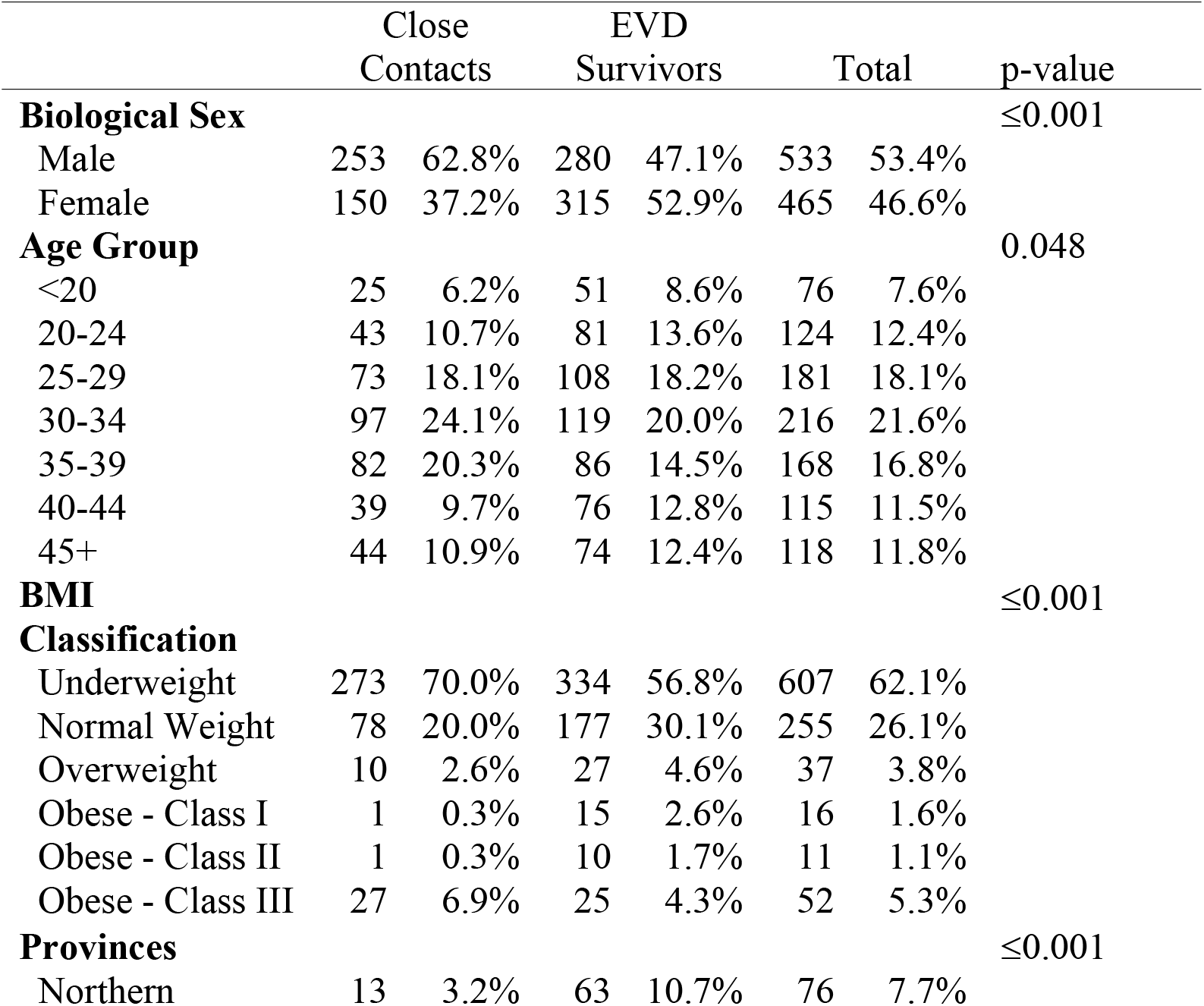
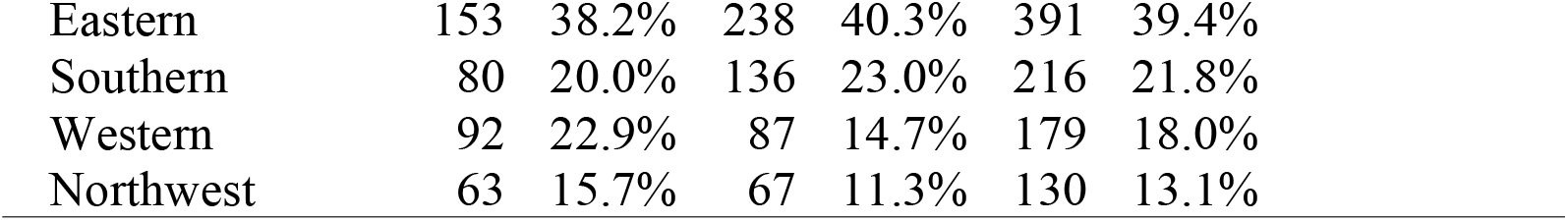
Demographic characteristics of participants by close contacts and EVD survivors.

|  | Close<br>Contacts |  | EVD<br>Survivors |  | Total |  | p-value |
| --- | --- | --- | --- | --- | --- | --- | --- |
| **Biological Sex** | | | | | | | $\leq 0.001$ |
| Male | 253 | 62.8% | 280 | 47.1% | 533 | 53.4% |  |
| Female | 150 | 37.2% | 315 | 52.9% | 465 | 46.6% |  |
| **Age Group** |  |  |  |  |  |  | 0.048 |
| <20 | 25 | 6.2% | 51 | 8.6% | 76 | 7.6% |  |
| 20-24 | 43 | 10.7% | 81 | 13.6% | 124 | 12.4% |  |
| 25-29 | 73 | 18.1% | 108 | 18.2% | 181 | 18.1% |  |
| 30-34 | 97 | 24.1% | 119 | 20.0% | 216 | 21.6% |  |
| 35-39 | 82 | 20.3% | 86 | 14.5% | 168 | 16.8% |  |
| 40-44 | 39 | 9.7% | 76 | 12.8% | 115 | 11.5% |  |
| 45+ | 44 | 10.9% | 74 | 12.4% | 118 | 11.8% |  |
| **BMI**<br>**Classification** | | | | | | | $\leq 0.001$ |
| Underweight | 273 | 70.0% | 334 | 56.8% | 607 | 62.1% |  |
| Normal Weight | 78 | 20.0% | 177 | 30.1% | 255 | 26.1% |  |
| Overweight | 10 | 2.6% | 27 | 4.6% | 37 | 3.8% |  |
| Obese - Class I | 1 | 0.3% | 15 | 2.6% | 16 | 1.6% |  |
| Obese - Class II | 1 | 0.3% | 10 | 1.7% | 11 | 1.1% |  |
| Obese - Class III | 27 | 6.9% | 25 | 4.3% | 52 | 5.3% |  |
| **Provinces** | | | | | | | $\leq 0.001$ |
| Northern | 13 | 3.2% | 63 | 10.7% | 76 | 7.7% |  |

|  |  |  |  |  |  |  |
| --- | --- | --- | --- | --- | --- | --- |
| Eastern | 153 | 38.2% | 238 | 40.3% | 391 | 39.4% |
| Southern | 80 | 20.0% | 136 | 23.0% | 216 | 21.8% |
| Western | 92 | 22.9% | 87 | 14.7% | 179 | 18.0% |
| Northwest | 63 | 15.7% | 67 | 11.3% | 130 | 13.1% |

### Main outcomes

EVD survivors were more likely to self-report symptomology across all body systems compared to close contacts (table 2) when controlled for sex, age group, BMI, and province of residence six-to-eight years after recovery. The symptom groups with the strongest association at the six-to-eight-year follow-up (table 3) were ENT symptoms (AOR: 8.75, 95% CI: 5.63 – 13.60, p < 0.001), ocular symptoms (AOR: 7.18, 95% CI: 5.02 – 10.25, p < 0.001), dermatological symptoms (AOR: 4.16, 95% CI: 3.06 – 5.65, p < 0.001) and cardiovascular symptoms (AOR: 2.96, 95% CI: 2.12 – 4.13, p < 0.001).

**Table 2:** Current Dermatological issues of study population by EVD survival status.

|  | Close Contact |  | EVD Survivor |  | p-value |
| --- | --- | --- | --- | --- | --- |
| **Chronic Medical Conditions** | 59 | 14.6% | 162 | 27.2% | $\leq 0.001$ |
| **Infections** | 254 | 63.0% | 411 | 69.1% | 0.048 |
| **Ocular Symptoms** | 80 | 19.9% | 336 | 56.5% | $\leq 0.001$ |
| **Ear, Nose and Throat Symptoms** | 47 | 11.7% | 250 | 42.0% | $\leq 0.001$ |
| **Neurological Symptoms** | 235 | 58.3% | 428 | 71.9% | $\leq 0.001$ |
| **Constitutional Symptoms** | 253 | 62.8% | 415 | 69.7% | 0.024 |
| **Cardiovascular Symptoms** | 103 | 25.6% | 280 | 47.1% | $\leq 0.001$ |
| **Respiratory Symptoms** | 146 | 36.2% | 291 | 48.9% | $\leq 0.001$ |
| **Gastrointestinal Symptoms** | 97 | 24.1% | 247 | 41.5% | $\leq 0.001$ |
| **Genitourinary Symptoms** | 202 | 50.1% | 374 | 62.9% | $\leq 0.001$ |
| **Musculoskeletal Symptoms** | 116 | 28.8% | 257 | 43.2% | $\leq 0.001$ |
| **Dermatology Symptoms** | 130 | 32.3% | 366 | 61.5% | $\leq 0.001$ |

**Table 3:**
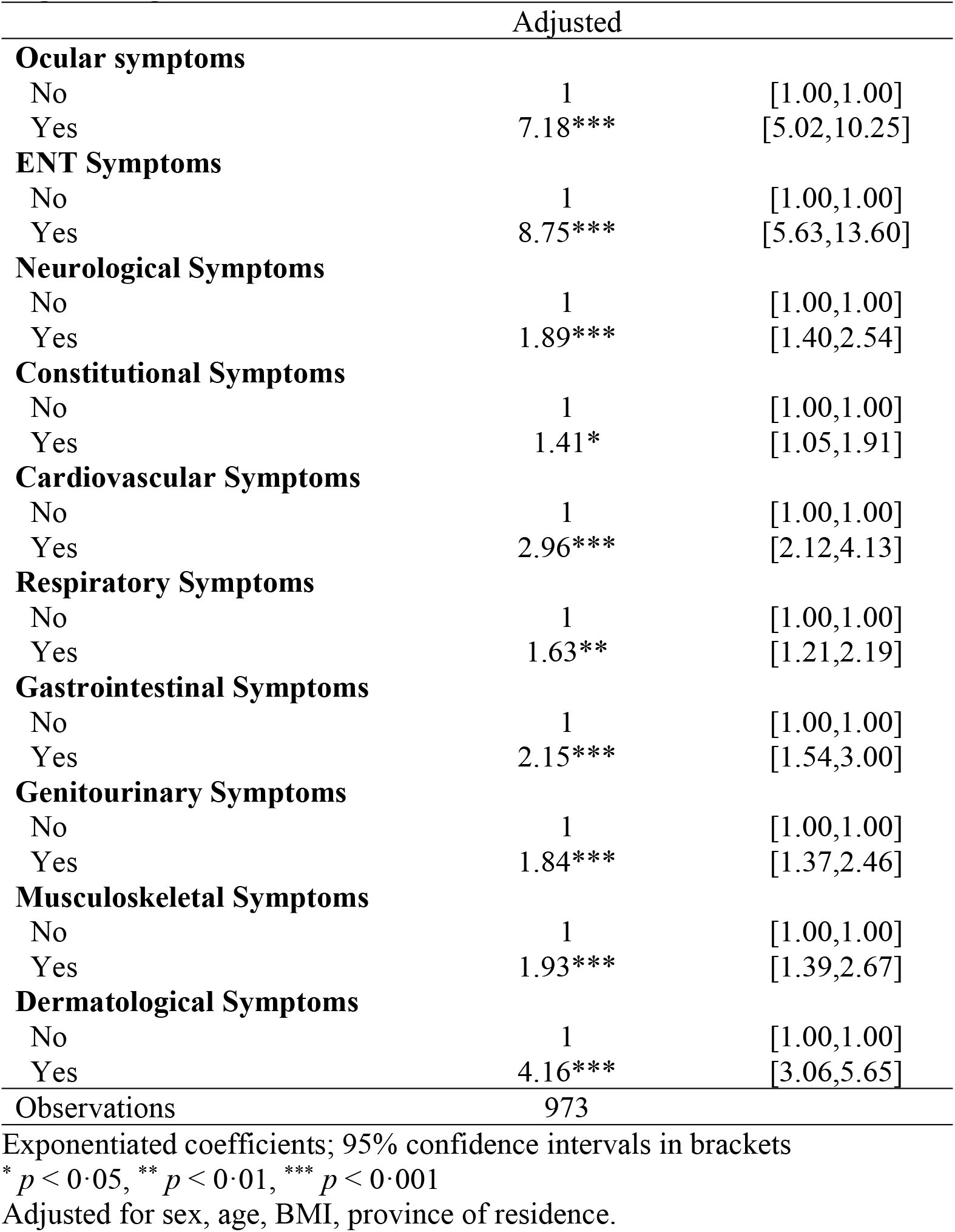
Associations with EVD survival by symptom group - Crude and Adjusted Odds Ratios (ORs) and 95% Confidence Intervals (95%CI), Logistic Regression Models.

### Commonly Reported Symptoms

ENT, Ocular, dermatological and cardiovascular symptoms were more prevalent in EVD survivors compared to close contacts. Frequencies of symptoms by symptom group are reported in Tables A-D of the appendix. Logistic regression adjusting for sex, age group, BMI, and province of residence was performed on major symptoms reported (Table 4). AORs for ENT symptoms (frequencies outlined in table A of the appendix) that were highly prevalent among survivors compared to close contacts were 7.18 for reduced hearing (95% CI: 5.02 – 10.25, p < 0.001) and 8.75 for otalgia (95% CI: 5.63 – 13.60, p < 0.001). Eye pain (AOR: 1.89, 95% CI: 1.40 – 2.54, p < 0.001), itchy eyes (AOR: 1.41, 95% CI: 1.05 – 1.91, p < 0.05) and eye redness (AOR: 2.96, 95% CI: 2.12 – 4.13, p < 0.001) were prevalent ocular symptomologies (frequencies outlined in table B of the appendix). The most prevalent dermatological symptoms (frequencies outlined in table C of the appendix) were itchy skin (AOR: 1.63, 95% CI: 1.21 – 2.19, p < 0.01) rash (AOR: 2.15, 95% CI: 1.54 – 3.00, p < 0.001) alopecia (AOR: 1.84, 95% CI: 1.37 – 2.46, p < 0.001) and change in skin colour (AOR: 1.93, 95% CI: 1.39 – 2.67, p < 0.001). The most reported cardiovascular symptoms (frequencies outlined in table D of the appendix) were chest pain (AOR: 5.39, 95% CI: 3.29 – 8.82, p < 0.001) and leg pain (AOR: 4.16, 95% CI: 3.06 – 5.65, p < 0.001).

**Table 4:** Associations with EVD survival by symptom – Crude and Adjusted Odds Ratios (AORs) and 95% Confidence Intervals (95%CI), Logistic Regression Models.

|  | AOR |  |
| --- | --- | --- |
| **Reduced hearing** |  |  |
| No | 1 | [1.00,1.00] |
| Yes | 7.18*** | [5.02,10.25] |
| **Pain in ears** |  |  |
| No | 1 | [1.00,1.00] |
| Yes | 8.75*** | [5.63,13.60] |
| **Eye pain** |  |  |
| No | 1 | [1.00,1.00] |
| Yes | 1.89*** | [1.40,2.54] |
| **Itchy eyes** |  |  |
| No | 1 | [1.00,1.00] |
| Yes | 1.41* | [1.05,1.91] |
| **Eye redness** |  |  |
| No | 1 | [1.00,1.00] |
| Yes | 2.96*** | [2.12,4.13] |
| **Itchy skin** |  |  |
| No | 1 | [1.00,1.00] |
| Yes | 1.63** | [1.21,2.19] |
| **Rash** |  |  |
| No | 1 | [1.00,1.00] |
| Yes | 2.15*** | [1.54,3.00] |
| **Alopecia** |  |  |
| No | 1 | [1.00,1.00] |
| Yes | 1.84*** | [1.37,2.46] |
| **Change in skin color** |  |  |
| No | 1 | [1.00,1.00] |
| Yes | 1.93*** | [1.39,2.67] |
| **Chest pain** |  |  |
| No | 1 | [1.00,1.00] |
| Yes | 5.39*** | [3.29,8.82] |
| **Leg pain** |  |  |
| No | 1 | [1.00,1.00] |
| Yes | 4.16*** | [3.06,5.65] |
| Observations | 973 |  |

### Reproductive Health Assessment

Amongst female participants, significantly more EVD survivors (n=56, 19.7%, p≤0.001) expressed decreased interest in sexual intercourse (table E appendix) than close contacts (n=6, 5.0%). The same was not observed for male EVD survivors (n=24, 9.3%, p≤0.086) and close contacts (n=11, 5.2%). Logistic regression analysis was performed for reproductive health (table 5 and 6) under the same conditions as described in earlier sections, stratified by sex at birth. EVD survival was significantly associated with several reproductive health variables, including sexual intercourse (AOR: 5.53, 95% CI: 3.04 – 10.07), decreased interest in sexual intercourse (AOR: 3.64, 95% CI: 1.34 – 9.86), irregular periods (AOR: 3.07, 95% CI: 1.75 – 5.36) and no recent pregnancies (AOR: 3.53, 95% CI: 1.69 – 7.34). For males, EVD survivorship was associated with reporting no conceived pregnancies in the last year (AOR: 2.73, 95% CI: 1.29-5.81).

**Table 5:** Associations Female Reproductive and Sexual Health Associations with EVD survival – Crude and Adjusted Odds Ratios (AORs) and 95% Confidence Intervals (95%CI), Logistic Regression Models.

|  | AOR |  |
| --- | --- | --- |
| **Sexual Intercourse in the Last Year** |  |  |
| No | 5.53*** | [3.04,10.07] |
| Yes | 1 | [1.00,1.00] |
| **Decreased Interest in Sexual Intercourse** |  |  |
| No | 1 | [1.00,1.00] |
| Yes | 3.64* | [1.34,9.86] |
| **Menstruation in the Last Year** |  |  |
| Regular Period | 1 | [1.00,1.00] |
| Irregular Period | 3.07*** | [1.75,5.36] |
| No Period | 1.56 | [0.57,4.27] |
| **Pregnant in the Last Year** |  |  |
| No | 3.53*** | [1.69,7.34] |
| Yes | 1 | [1.00,1.00] |
| **Given Birth** |  |  |
| No | 1 | [1.00,1.00] |
| Yes | 1.28 | [0.74,2.20] |
| Observations | 384 |  |

Exponentiated coefficients; 95% confidence intervals in brackets
\* $p < 0.05$ , \*\* $p < 0.01$ , \*\*\* $p < 0.001$
Adjusted for sex, age, BMI, province of residence.

**Table 6:**
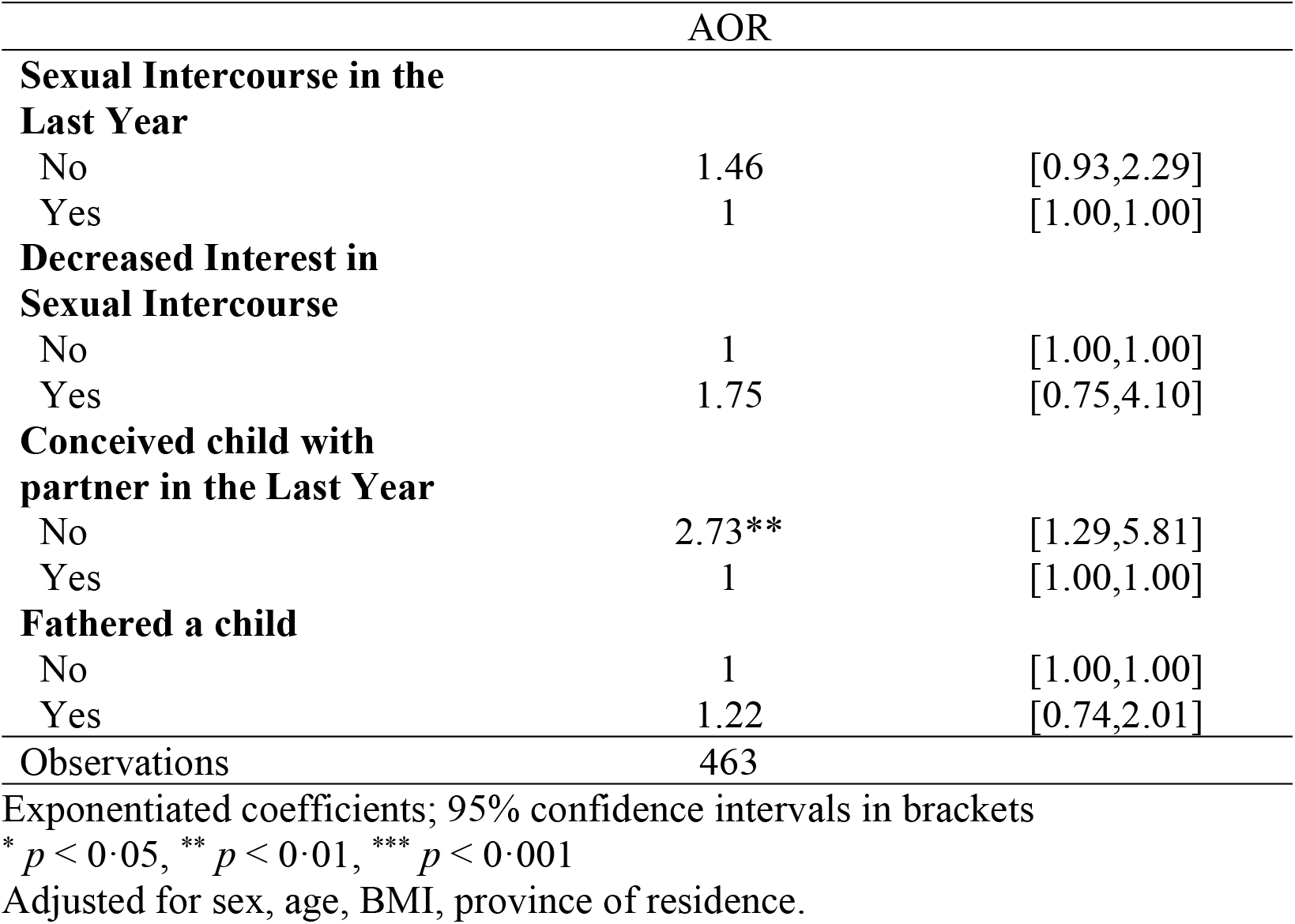
Associations Male Reproductive and Sexual Health Associations with EVD survival – Crude and Adjusted Odds Ratios (AORs) and 95% Confidence Intervals (95%CI), Logistic Regression Models.

## Discussion

In this study, we present the most prevalent clinical symptoms experienced by EVD survivors compared to their close contacts, 6-8 years after the West Africa EVD outbreak. Other studies have reported on persistent ocular manifestations in survivors dating to the Kikwit outbreak, where a two year follow-up study identified ocular pain, blurry vision and light sensitivity[24]. The same study also identified the presence of highly prevalent symptoms including joint pain, hearing loss and fatigue[24]. These symptoms continue to be among the most reported sequelae in available literature and were among the most reported in this study[3, 4, 6, 7, 14, 16-25]. There are several reports on clinical health outcomes in the immediate aftermath of the West African EVD epidemic, however, there has been a paucity of information on the longer-term clinical health outcomes of EVD survivors.

While it has previously been reported that persistence of sequelae diminishes over time, our results suggest that in Sierra Leone this has not been the case[5, 7, 16] (table 2). We report on five symptom types reported by >50% of EVD survivors 6-8 years post recovery, and all but one reported by >40% of EVD survivors[5]. Recent studies including by Wohl et al., the only other published study with long term follow-up of survivors in Liberia included a study population of 326 participants who were followed up to five years from Ebola treatment Center (ETC) discharge[7]. It is difficult to compare the findings between two studies in different countries utilizing different survey tools, however some of the differences are notable in results at 5+ years post recovery; including fatigue (12.2% vs 20.2% reported in table I of appendix), headache (20.3% vs 64.4% in table H of appendix), hearing loss (2.8% vs 13.9% in table A of appendix), joint pain (15.8% vs 26.7% in table J of appendix), muscle pain (0.0% vs 5.5% in table J of appendix) and vision impairment (17.4% vs 8.1% in table B of appendix). The experiences reported by survivors of the same epidemic in a similar time frame of assessment demonstrate the variable experiences of long-term sequelae by survivors based on their geographic location and environment. It is challenging to hypothesize on which factors may account for the differences observed in the rates of these various symptomologies (As listed in table 2); however, population sample size, living conditions and health care access are plausible factors that may affect these differences observed. When comparing symptomology to reported rates from another recent study conducted by Bond et al. in Eastern Sierra Leone, with 375 EVD survivors and 1040 close contacts, similar findings of physical symptoms were significantly more commonly reported by survivors compared with close contacts[17]. This study was conducted between 1.6 and 3.3 years after discharge from ETCs suggesting that symptomology rates have remained elevated and stable over the subsequent 3-5 years from the initial recovery period to the 6–8-year follow-up time point reported in our study. Similar findings to those presented here have previously been observed in West Africa regarding reproductive health of female and male EVD survivors. A study conducted from January to April 2016 in Montserrado county, Liberia found that 16.2% of male EVD survivors surveyed reported testicular pain or erectile dysfunction, while 19.7% of female EVD survivors reported having menstrual problems including amenorrhoea and other menstrual irregularities[14]. Another study conducted in February 2015 in Bombali District, Sierra Leone found that 2.2% of female EVD survivors reported a menstrual disorder and 3.4% of male EVD survivors reported erectile dysfunction[29]. A study conducted in Liberia between 2016 and 2017 including 111 female participants from Monrovia, reported 25.7% of female participants had irregular menstruation of unknown cause[30]. The same group reported in a qualitative study including 69 women, that 50% of female survivors from Liberia had reported decreased sexual libido following recovery with several women also reporting pain during intercourse, and 46% of participants reporting some of menstrual irregularity[31]. We made similar observations in our study (table 6 and tables E and F of the appendix,) with female survivors also reporting menstrual changes and changes in sexual libido. Reproductive health changes experienced by EVD survivors remain understudied. Additionally, there are long-term social and mental health effects, as we have previously reported in this cohort[26, 32]. Many survivors experience social isolation, with their relationship status being directly associated with stigmatization. The mental health effects are also profound, with 45.7% of EVD survivors reporting post-traumatic stress disorder related to their Ebola experience and its impact on their interpersonal relationships[26, 32]. There is a lack of data on the factors related to the host and the disease that predict symptoms and the mechanisms behind their persistence. Understanding the viral and immune-related mediators may help develop targeted treatment plans to address these chronic symptoms for EVD survivors[24, 33].

Investigations into the mechanisms that explain the sequelae reported by EVD survivors have been limited. However, some studies have found associations with various symptoms[24, 33]. A 23-month follow-up study identified increased blood markers of inflammation, intestinal tissue damage, T-cell and B-cell activation and a depletion of circulating dendritic cells[9]. Additionally the level of adaptive immune cells remained highly elevated two years after recovery in EVD survivors tested[9]. The study also found that immune cell phenotypes shifted towards activated cytotoxic T cells, exhausted B cells, proinflammatory monocytes and non-classical NK cells, which are common in chronic viral infections. Elevated levels of inflammatory markers and activation of antiviral pathways have been reported to persist in the absence of detectable virus[9]. Specifically D dimer, IFN-γ, IL-8 and TNF-alpha were shown to persist. Markers of chronic immune activation such as CCL5 and sCD40L, and markers of potential liver and kidney damage, including higher levels of alanine transaminase, aspartate transaminase, bilirubin, creatine and creatine kinase, were also found to persist[6, 9, 34]. These findings demonstrate that the chronic effects of EVD infection continue to manifest in physical symptoms that severely impact the day-to-day lives of EVD survivors across all organ systems.

Symptoms have made it difficult to get back to their prior lives they lived[14]. Among the most reported symptoms in this study were dermatological issues, including itchy skin, rashes, alopecia, and hyperpigmentation. While these symptoms have been previously reported, there are no recent reports discussing them in detail[3, 14]. Given their high visibility and prevalence (61.5% in this cohort and previously reported at 48.8%), these symptoms can be highly discriminating and visible sequelae among survivors[14]. Previous reports suggest reintegration into the community was limited[20], with many survivors reporting a lower socioeconomic status than prior to EVD (90%), less favourable work (79%) and worse psychological status (60%)[20]. In Kikwit, Democratic Republic of the Congo 70% of survivors indicated capacity to work was worse 21 months following infection compared to before suggesting that the physical sequalae, as observed in this study affects their ability to return to their prior lives[24]. 60.9% of women seeking care at the survivor clinic in Kenema were female[22]. Some survivors may benefit from rheumatologic, musculoskeletal, or neurologic evaluation in the recovery phase[11].

A limitation of this study is the reliance on self-reporting symptomologies without formal assessment of impairments through physical examinations. This was mitigated by using validated standardized questionnaires with objective measurements of symptoms and direct comparisons to a control population. The study’s cross-sectional design limited our ability to establish cause- and-effect relationships or analyse change over time meaningfully. Recruiting close contacts as a control population allowed us to match the economic, geographic and social experiences between the two study groups as much as possible. Exploratory analyses conducted may have increased the risk of type I error in this study. It is possible that individuals with more severe symptoms were more likely to participate, but this was mitigated through broad recruitment both in terms of the number of survivors and their geographic distribution.

## Conclusions

Our results indicate that many, if not most, EVD survivors from the West Africa EVD epidemic continue to experience persistent health issues. Factors such as inconsistent follow up on the management of sequelae[11], limited access to healthcare services difficulty in obtaining necessary mediations or follow up care, poor health seeking behavior, and inadequate access to quality healthcare have compounded the challenges in managing these symptoms.[35] Additionally, the withdrawal of support without a proper transition period has led to issues like food insecurity, further exacerbating the long-term sequelae observed in our study.

Given the chronic nature of EVD-associated sequelae, it is crucial to consider funding and strengthening existing medical infrastructure in affected countries to ensure long-term follow up and care.[25]. Sustained support for survivor services in West Africa is essential, and funding for survivor follow-up should be integrated into future outbreak response planning planning provide to provide adequate care both during acute infection and throughout the post-recovery period[25].

## Data Availability

Due to the sensitive nature of our survey and the involvement of human participants we did not receive approval to collect raw participant identified data from The University of Manitoba Research Ethics Board or the Sierra Leone Ethics and Scientific Review Committee. While participants additionally consented to the study with the knowledge that their data was collected in an anonymized manner with the only record of participants being record of payment only accessible by JK and JBK and securely stored in an encrypted file. Upon reasonable request however, the anonymized raw study data can be made available through the following persons 1) from UM REB by contacting the office 2) Edward Foday, Research and Publications Specialist, Sierra Leone Ethics and Scientific Review Committee, Directorate of Policy, Planning and Information, Ministry of Health and Sanitation, Fifth Floor, Youyi Building, East Wing, Freetown, Sierra Leone.

## Contributors

JK and JBK conceived the study, while BGS, JBK, SYS, and JK designed the study. JBK lead the recruitment and in-country support in Sierra Leone. BGS BKT SYS analysed the data. BGS prepared the manuscript. JK supervised the study, analysis and writing process. All authors contributed to the intellectual content of the manuscript and read and approved the final version of the manuscript.

## Declaration of Interest

The authors declare no competing interests.

### Data Sharing

Due to the sensitive nature of our survey and the involvement of human participants we did not receive approval to collect raw participant identified data from The University of Manitoba Research Ethics Board or the Sierra Leone Ethics and Scientific Review Committee. While participants additionally consented to the study with the knowledge that their data was collected in an anonymized manner with the only record of participants being record of payment only accessible by JK and JBK and securely stored in an encrypted file. Upon reasonable request however, the anonymized raw study data can be made available through the following persons; 1) from UM REB by contacting the office 2) Edward Foday, Research and Publications Specialist, Sierra Leone Ethics and Scientific Review Committee, Directorate of Policy, Planning and Information, Ministry of Health and Sanitation, Fifth Floor, Youyi Building, East Wing, Freetown, Sierra Leone.

## Acknowledgements

We would like to thank our funding sources JK is funded by a Tier 2 Canada Research Chair in the Molecular Pathogenesis of Emerging and Re-Emerging Viruses provided by the Canadian Institutes of Health Research (Grant no. 950-231498), and the Canadian Institutes of Health Research (Grant no. PJT-175098). BKT has received funding support from NIH Building Interdisciplinary Careers in Women’s Health (BIRCWH) Grant (K12HD085850) and from Emory’s Center AIDS Research Grant (P30 AI050409). We would like to thank both the EVD survivor and close contact participants of this study. We would also like to thank the participant recruiters for their work and support. We would also like to thank the Sierra Leone Ebola Survivors Association.

## Abbreviations

EVD: (Ebola virus disease)
BMI: (body mass index)
ENT: (ear, nose and throat)
FSFI: (Female sexual function index)
FSAD: (Female sexual arousal disorder)
IIEF: (International index of erectile function)
IQR: (Interquartile range)
AOR: (Adjusted odds ratio)
CI: (Confidence interval)
ETC: (Ebola treatment Centre)

